# Impaired Humoral Immunity to SARS-CoV-2 Vaccination in Non-Hodgkin Lymphoma and CLL Patients

**DOI:** 10.1101/2021.06.02.21257804

**Authors:** Catherine Diefenbach, Jessica Caro, Akiko Koide, Michael Grossbard, Judith D. Goldberg, Bruce Raphael, Kenneth Hymes, Tibor Moskovits, Maxim Kreditor, David Kaminetzky, Shella Saint Fleur-Lominy, Jun Choi, Sara A. Thannickal, Kenneth A. Stapleford, Shohei Koide

## Abstract

Patients with hematologic malignancies are a high priority for SARS-CoV-2 vaccination, yet the benefit they will derive is uncertain. We investigated the humoral response to vaccination in 53 non-Hodgkin lymphoma (NHL), Hodgkin lymphoma (HL), or CLL patients. Peripheral blood was obtained 2 weeks after first vaccination and 6 weeks after second vaccination for antibody profiling using the multiplex bead-binding assay. Serum IgG, IgA, and IgM antibody levels to the spike specific receptor binding domain (RBD) were evaluated as a measure of response. Subsequently, antibody-positive serum were assayed for neutralization capacity against authentic SARS-CoV-2. Histology was 68% lymphoma and 32% CLL; groups were: patients receiving anti-CD20-based therapy (45%), monitored with disease (28%), receiving BTK inhibitors (19%), or chemotherapy (all HL) (8%). SARS-CoV-2 specific RBD IgG antibody response was decreased across all NHL and CLL groups: 25%, 73%, and 40%, respectively. Antibody IgG titers were significantly reduced (p < 0.001) for CD20 treated and targeted therapy patients, and (p = 0.003) for monitored patients. In 94% of patients evaluated after first and second vaccination, antibody titers did not significantly boost after second vaccination. Only 13% of CD20 treated and 13% of monitored patients generated neutralizing antibodies to SARS-CoV-2 with ICD50s 135 to 1767, and 445 and > 10240. This data has profound implications given the current guidance relaxing masking restrictions and for timing of vaccinations. Unless immunity is confirmed with laboratory testing, these patients should continue to mask, socially distance, and to avoid close contact with non-vaccinated individuals.

**Statement of Translational Relevance:** Non Hodgkin lymphoma (NHL) and Chronic Lymphocytic leukemia (CLL) patients who are treated with anti-CD20 antibody therapy, BTK inhibitor therapy, or who are monitored with active disease, have decreased antibody response to SARS-CoV-2 vaccination and decreased antibody titers compared to healthy controls. Antibody titers do not boost following second vaccination, and very few patients generate neutralizing antibodies against SARS-CoV-2. This data is of particular importance, given the recent guidance from the CDC that vaccinated patients no longer need to be masked indoors as well as outdoors. Patients with NHL or CLL who fall into these categories should not consider their immunity from vaccination to be assured. If infected with SARS-CoV-2, they should be a high priority for monoclonal antibody directed therapy. Unless immune response to vaccination is confirmed with laboratory testing, they should continue to mask, socially distance, and to avoid close contact with non-vaccinated individuals.

## Introduction

Patients with hematologic malignancies are among the highest risk groups for poor outcomes to SARS-CoV-2 due to immune compromise from their disease and concurrent anti-cancer therapy. A study of cancer patients at a NYC hospital found that hematologic malignancies, compared to solid cancers, were associated with a higher rate of mortality (37% vs. 25%) and ICU admission (26% vs. 19%) (1). A larger retrospective UK study of 1044 cancer patients with SARS-CoV-2 infection had similar findings, with patients with hematologic malignancies having a more severe trajectory than patients with solid organ transplant (2). This vulnerable population is a high priority for vaccination, yet the benefit they will derive from vaccination is uncertain. Lymphoma and chronic lymphocytic leukemia (CLL) patients have a reduced humoral response to influenza and pneumococcal vaccines, including a poor response to recall antigens in Hodgkin lymphoma (HL) patients (3-6). Although Phase 3 registration trials of all 3 FDA approved SARS-CoV-2 vaccines showed high efficacy in preventing symptomatic infection independent of age, none of them included patients with hematologic malignancies (7-9). Emerging data suggest that CLL patients have a reduced humoral immune response to SARS-CoV-2 vaccination (10, 11). This has profound implications for the safety of these patients, given the current guidance relaxing masking restrictions for vaccinated patients. Here, we investigated prospectively the question of whether in a cohort of non-Hodgkin lymphoma (NHL), HL, and CLL patients demonstrate impaired humoral immunity to SARS-CoV-2 vaccination at NYU Langone Health Perlmutter Cancer Center (PCC) hospitals.

## Methods

The Institutional Review Board of the Perlmutter Cancer Center at NYU Langone Health reviewed and approved the study, which was conducted according to the Declaration of Helsinki and International Harmonization Guidelines for Good Practice. All patients signed informed consent for the study. We evaluated humoral response to vaccination in patients with NHL, HL, or CLL who were at least 18 years of age and were actively receiving anti-lymphoma treatment, were within 6 months of receiving therapy, or were monitored expectantly with active disease. All patients who were within 12 weeks of SARS-CoV-2 vaccination were eligible. Peripheral blood (40mL per time-point) was obtained at 2 weeks (+/-1 week) after the first vaccination and at 6 weeks (+/-2 weeks) after the second or single dose vaccination. All blood was processed within 24 hours of collection for peripheral blood mononuclear cells (PBMCs) and plasma. When patients were consented prior to vaccination, a pre-vaccine baseline blood draw was obtained. For those patients who completed the second vaccine, post vaccination levels were compared for those patients who completed two vaccinations. For patients who completed one vaccination, the post first vaccination time-point is used, for patients who completed both vaccinations, the post second vaccination time-point was used. Differences between these values were compared among patients with both time points. Absolute lymphocyte count (ALC) was extracted from the electronic medical record (EMR) from clinic visit immediately prior to SARS-CoV-2 vaccination.

Antibody profiling was performed using the multiplex bead-binding assay, as previously described (12, 13). Briefly, biotinylated spike and RBD proteins produced in house and biotinylated nucleocapsid protein purchased from Sino Biological (catalog number 40588-V27B-B) were immobilized on the MultiCyt® QBeads® Streptavidin Coated panel QSAv1,2 and 3 (Sartorius catalog number 90792) and incubated with plasma samples diluted in PBS with 0.1 % Tween 20 and 1% skim milk (Millipore Sigma catalog number 1.15363.0500). We detected bound IgG, IgA and IgM using anti-human IgG-Alexa 488 (Jackson Immunoresearch catalog number 109-545-098), anti-human IgA-PE (Jackson Immunoresearch; catalog number 109-115-011) and anti-human IgM-DyLight405 (Jackson Immunoresearch; catalog number 709-475-073), diluted in PBS with 0.1 % Tween 20 and 1 % BSA to 1:800, 1:100 and 1:200 respectively, on a Yeti ZE5 Cell Analyzer (Bio-Rad) and analyzed the data using FlowJo (BD, version 10.7.1). Cutoff values were defined as the mean plus 3 x s.d. of the median fluorescence intensities from six pre-COVID samples (negative controls) described previously (14).

For virus neutralization assays, icSARS-CoV-2-mNG (isolate USA/WA/1/2020 expressing a mNeon Green reporter), was obtained from the UTMB World Reference Center for Emerging Viruses and Arboviruses(15) and amplified once in Vero E6 cells (ATCC CRL-1586) to generated a working stock. We seeded 20,000 Vero E6 cells/well in 96 well plates the day before infection. Patient serum was 2-fold serially diluted (ranging from 1:20 to 1:10,240) in DMEM supplemented with 1% nonessential amino acids, 10 mM HEPES, and 2% fetal bovine serum. Each serum dilution was mixed 1:1 (vol/vol) with 5000 PFU of SARS-CoV-2-mNG and incubated at 37°C for 1 hr. The virus:serum mix was then added to the Vero E6 cell containing plates and incubated at 37°C for 24 hrs. After incubation, cells were fixed with 10% formalin, stained with DAPI, and virus positive cells were quantified using a CellInsight CX7 High-content microscope using a cut-off of three standard deviations from negative to be scored as an infected cell.

For each of the individual immunoglobulin measurements, mean levels for each group of patients were compared with the levels of the 3 healthy vaccinated controls using 2-sample t-tests. A two-sample 2-sided t-test with alpha=0.05 was used to compare ALC between patient subgroups and unvaccinated controls with adjustment for multiple comparisons. To adjust for the 5 multiple comparisons with the 3 healthy controls for each immunoglobulin individually, we use a Bonferroni correction and consider differences to be statistically significant if p is less than 0.01 (0.05/5).

### Data Sharing

Contact the corresponding author for original data

## Results

### Patient Demographic Characteristics

The analysis included 53 patients with diagnosis of NHL, HL, or CLL, who were actively receiving anti-lymphoma treatment, were within six months of receiving therapy, or were monitored expectantly with active disease. Demographic characteristics are shown in **Table 1**. The median age was 63, and patients were evenly divided by gender. All 53 patients received SARS-CoV-2 vaccination; 52 patients had blood draws after their first vaccination (visit 1) and 18 of these 52 patients had blood draws after their second vaccination (visit 2), one patient only had blood drawn after their second vaccination. Lymphoma patients were 68% of the population and CLL 32%. The most common histologies were CLL/small lymphocytic lymphoma (SLL) in 17 (32%) and diffuse large B cell lymphoma (DLBCL) in 10 (19%). Other histologies are shown in **Table 1**. Three patients who only had pre-vaccine blood draw were not included in the analysis of vaccine immunogenicity but served as negative controls. Three healthy vaccinated individuals and two convalescent vaccinated individuals were used as positive controls. Twenty-four patients (45%) received anti-CD20-based therapy and 15 patients (28%) were monitored with active disease, of these 10 were treatment naïve and 5 had received previous treatment with a mean duration from treatment to vaccination of 1671 days. Ten patients (19%) received targeted therapy (all BTK inhibitors), and 4 patients (8%) with HL received chemotherapy with adriamycin, bleomycin, vinblastine, dacarbazine (ABVD). Forty-one patients (77%) received the Pfizer vaccine, and 12 (23%) received the Moderna vaccine. Thirty patients (57%) were vaccinated during treatment, 6 patients (11%) received treatment within 6 months of vaccination, and 2 patients (4%) received treatment after their first vaccination. The median time between therapy and vaccination was 9.5 days (range 0-152) for all therapy and 42.5 days (range 3-285) for anti-CD20 therapy.

**Table 1:** Patient Demographic Characteristics.

|  | Overall<br>(n = 53) | CD-20 based<br>therapy<br>(n = 24) | Chemothera<br>py (n = 4) | Monitoring<br>(n = 15) | Targeted<br>therapy<br>(n = 10) |
| --- | --- | --- | --- | --- | --- |
| **Age, years** |  |  |  |  |  |
| Median (range) | 63 (25-98) | 64 (30-98) | 31 (25-35) | 62 (45-93) | 63 (42-75) |
| **Gender** |  |  |  |  |  |
| Female | 25 (47%) | 13 (54%) | 3 (75%) | 8 (53%) | 1 (10%) |
| **Histologic diagnosis** |  |  |  |  |  |
| CLL/SLL | 17 (32%) | 2 (8%) | 0 (0%) | 8 (53%) | 7 (70%) |
| Hodgkin lymphoma | 4 (8%) | 0 (0%) | 4 (100%) | 0 (0%) | 0 (0%) |
| DLBCL | 10 (19%) | 7 (29%) | 0 (0%) | 2 (13%) | 1 (10%) |
| Follicular lymphoma | 7 (13%) | 7 (29%) | 0 (0%) | 0 (0%) | 0 (0%) |
| Mantle cell lymphoma | 3 (6%) | 2 (8%) | 0 (0%) | 1 (7%) | 0 (0%) |
| Marginal zone lymphoma | 7 (13%) | 4 (17%) | 0 (0%) | 3 (20%) | 0 (0%) |
| Low-grade B cell lymphoma | 1 (2%) | 1 (4%) | 0 (0%) | 0 (0%) | 0 (0%) |
| Burkitt lymphoma | 1 (2%) | 1 (4%) | 0 (0%) | 0 (0%) | 0 (0%) |
| Waldenstrom's<br>macroglobulinemia | 3 (6%) | 0 (0%) | 0 (0%) | 1 (7%) | 2 (20%) |
| **Type of COVID-19 vaccine** |  |  |  |  |  |
| Pfizer | 41 (77%) | 19 (79%) | 4 (100%) | 9 (60%) | 9 (90%) |
| Moderna | 12 (23%) | 5 (21%) | 0 (0%) | 6 (40%) | 1 (10%) |
| **Status of Treatment** |  |  |  |  |  |
| Treatment Pre-vaccine | 6 (11%) | 5 (21%) | 1 (25%) | 0 (0%) | 0 (0%) |
| Treatment Post-vaccine | 2 (4%) | 2 (8.3%) | 0 (0%) | 0 (0%) | 0 (0%) |
| Pre- and post-vaccine | 30 (57%) | 17 (71%) | 3 (75%) | 0 (0%) | 10 (100%) |
| No Treatment (Monitored) | 15 (28%) | 0 (0%) | 0 (0%) | 15 (100%) | 0 (0%) |
| **Status of vaccination** |  |  |  |  |  |
| Evaluated after first vaccine | 52 (98%) | 23 (96%) | 4 (100%) | 15 (100%) | 10 (100%) |
| Evaluated after second vaccine | 19 (36%) | 12 (50%) | 1 (25%) | 1 (7%) | 5 (50%) |
| Evaluated after both vaccines | 18 (34%) | 11 (46%) | 1 (25%) | 1 (7%) | 5 (50%) |
| Evaluated after second vaccine<br>only | 1 (2%) | 1 (2%) | 0 (0%) | 0 (0%) | 0 (0%) |
| **Days from Therapy to First**<br>**Vaccination** |  |  |  |  |  |
| Median | 9.5 | 42.5 | 17 | N/A | 0 |
| Range | 0 - 152 | 3 - 285 | 4 - 152 | N/A | 0 - 13 |
| **Monitored Patients** |  |  |  |  |  |
| Previously Treated |  |  |  | 5 (33%) |  |
| Never Treated |  |  |  | 10 (67%) |  |
| Days from Prior Therapy, mean (range) |  |  |  | 1671 (334-3924) |  |

### Decreased Antibody Response in NHL and CLL across all Histologies

The antibody mediated humoral response to the SARS-CoV-2 whole spike and spike receptor binding domain (RBD) was evaluated for 53 vaccinated patients. Antibody response to the RBD has been shown to be more selective to SARS-CoV-2 than that to the whole spike, because of lower sequence homology of the RBD among related viruses than the whole spike (16, 17). Accordingly, we consider antibody levels to the RBD as a measure of response to vaccination. Six (25%) of 24 patients receiving anti CD20 based therapy +/-chemotherapy exhibited an IgG immune response to the SARS-CoV-2 specific RBD antigen (**Figure 1, Table 2**). Antibody responses were higher in patients who were monitored expectantly with 73% exhibiting an IgG immune response to the RBD; one (7%) had a prior SARS-CoV-2 infection as demonstrated by positive antibody response to nucleocapsid (NP) IgG. Patients receiving BTK inhibitors (one of whom had a positive NP IgG) had 40% IgG antibody response to the RBD. In contrast, all HL patients receiving standard ABVD chemotherapy had a positive antibody response (100%) to SARS-CoV-2 specific RBD IgG without positivity for NP IgG **(Figure 1, Table 1)**. In addition, we quantified the amount of IgA and IgM to the spike and spike RBD and found that for all groups, antibody response was higher to the whole spike IgG than to the SARS-CoV-2 specific RBD, most likely reflecting the presence of preexisting immunity to related viruses (17) (**Table 2**). Twenty-five percent of patients receiving CD20 based therapy were positive for whole spike IgM and 46% were positive for whole spike IgA, however only 4% were positive for RBD IgG and 17% positive to RBD IgA. Patients being monitored had 40% positive antibody response to whole spike IgM and 53% to RBD IgG compared with 40% to RBD IgM and 25% to RBD IgA. In the BTK inhibitor therapy group 50% of patients had antibody response to whole spike IgM and 70% to whole spike IgA compared to 0% to RBD IgM and 30% to RBD IgA. The HL patients had the highest antibody response with 75% positive to whole spike IgM and 100% positive to whole spike IgA, and 50% positive to RBD IgM, and 100% positive to RBD IgA (**Figure 1, Table 2**).

**Figure 1.**
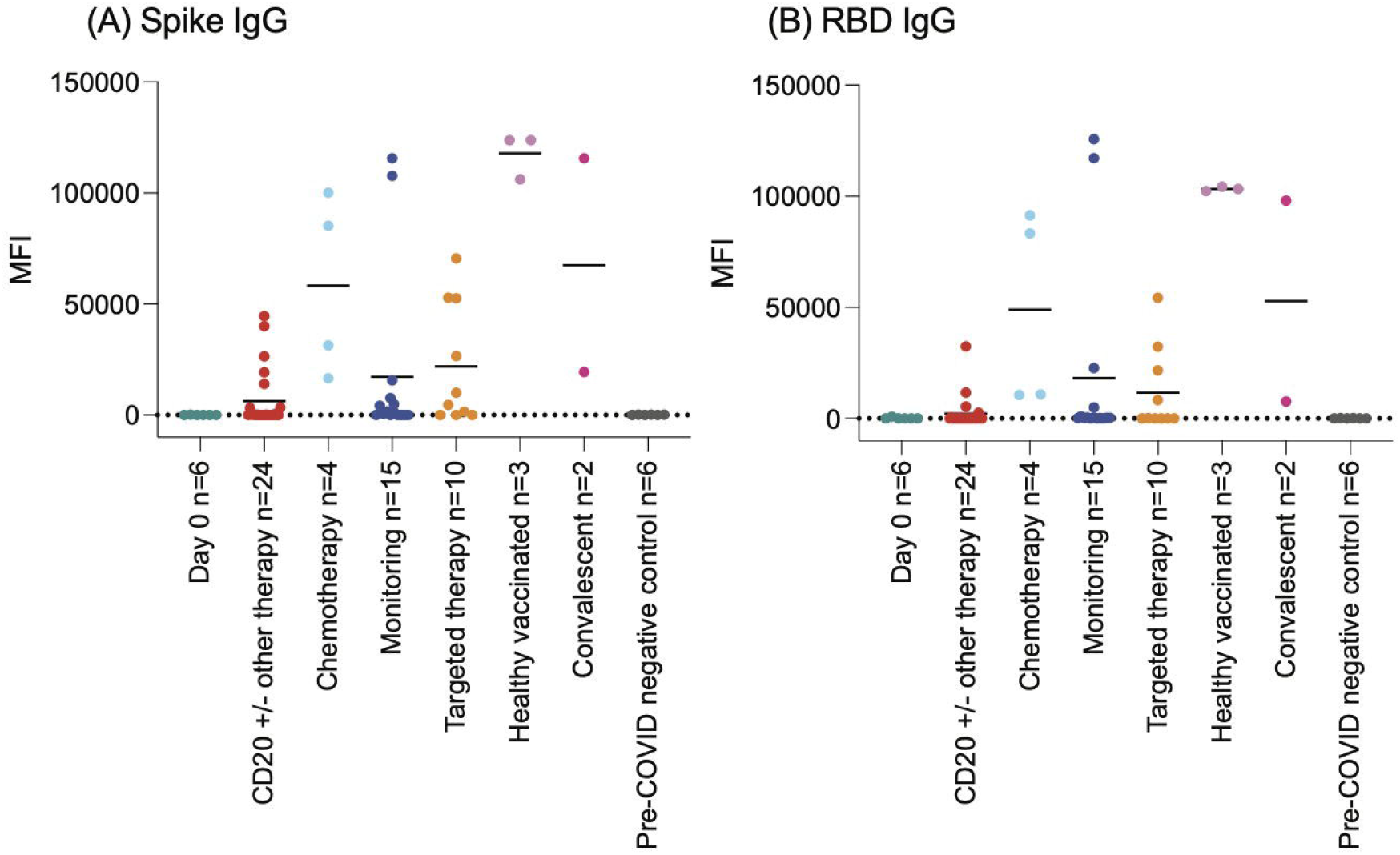
The levels of anti-Spike (A) and anti-RBD (B) IgG categorized by treatment. IgG levels measured in the median fluorescence intensity (MFI) are shown with bars indicating the mean of each group’s MFI.

**Table 2:** Incidence of Positive Immunoglobulin Levels for Spike IgG and RBD IgG.

| Patient Group | Number | Spike IgG | Spike IgM | Spike IgA | RBD IgG | RBD IgM | RBD IgA |
| --- | --- | --- | --- | --- | --- | --- | --- |
| Healthy vaccinated controls | 3 | 100% | 100% | 100% | 100% | 67% | 100% |
| Convalescent controls | 2 | 100% | 100% | 100% | 100% | 100% | 100% |
| CD20-based therapy | 24 | 38% | 25% | 46% | 25% | 4% | 17% |
| Chemotherapy | 4 | 100% | 75% | 100% | 100% | 50% | 100% |
| Monitoring | 15 | 60% | 40% | 53% | 73% | 40% | 25% |
| Targeted therapy | 10 | 70% | 50% | 70% | 40% | 3% | 30% |

### Decreased anti SARS-CoV-2 Antibody Titers in Antibody Positive Patients

To assess the quality of the antibody response, we evaluated antibody titers in patients who demonstrated a positive antibody response. For all NHL and CLL patients, antibody titers were significantly reduced compared to normal and controls **(Figure 1, Table 3)**. The mean RBD IgG level for healthy vaccinated controls was 1.03 × 10^5^ mg/dl. Ten antibody positive patients receiving CD20 antibody-based therapy had a mean IgG titer to RBD of 5.32 × 10^3^ mg/dl (p<0.0001). Eleven antibody positive patients in the monitoring group had a mean titer of 2.49 × 10^4^ mg/dl to RBD IgG (p = 0.0003). Patients receiving BTK inhibitors had mean RBD IgG of 1.67 × 10^4^ (p<0.0001). In contrast, the titers for the HL patients receiving standard ABVD chemotherapy were not significantly lower than control titers with mean RBD IgG of 4.90 × 10^4^ (p=0.09). There were no significant associations of higher antibody titers with age, NHL histology, type of vaccine (Pfizer vs. Moderna), or time from treatment.

**Table 3:**
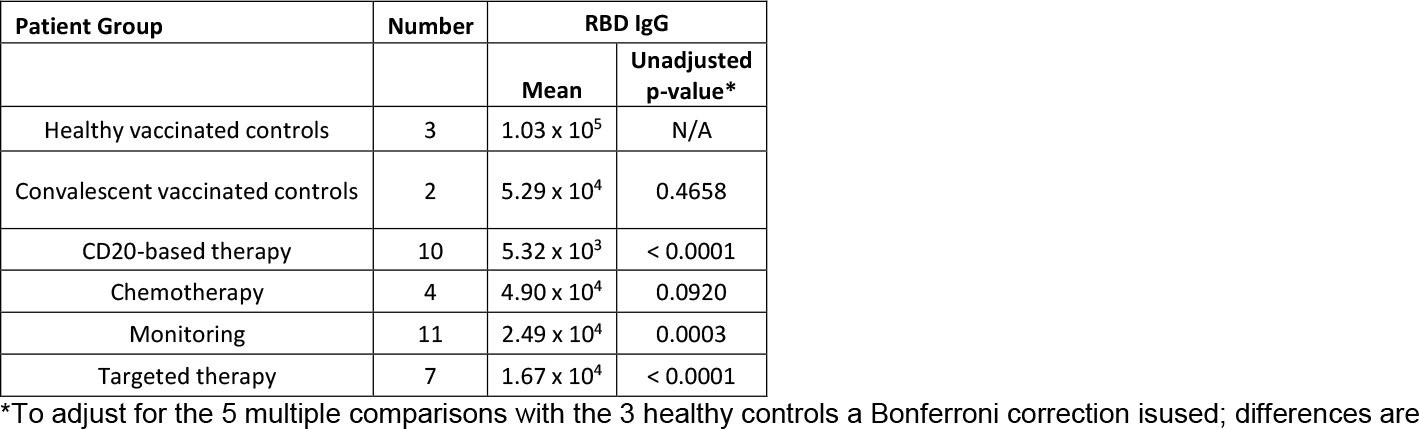
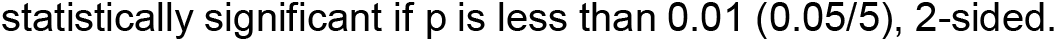
Mean Immunoglobulin Levels for Patients with Positive Spike IgG and/or RBD IgG TestedAgainst Vaccinated Controls Compared to Healthy Vaccinated Controls.

| Patient Group | Number | RBD IgG |  |
| --- | --- | --- | --- |
|  |  | Mean | Unadjusted p-value* |
| Healthy vaccinated controls | 3 | $1.03 \times 10^5$ | N/A |
| Convalescent vaccinated controls | 2 | $5.29 \times 10^4$ | 0.4658 |
| CD20-based therapy | 10 | $5.32 \times 10^3$ | < 0.0001 |
| Chemotherapy | 4 | $4.90 \times 10^4$ | 0.0920 |
| Monitoring | 11 | $2.49 \times 10^4$ | 0.0003 |
| Targeted therapy | 7 | $1.67 \times 10^4$ | < 0.0001 |

\*To adjust for the 5 multiple comparisons with the 3 healthy controls a Bonferroni correction is used; differences are
statistically significant if $p$ is less than 0.01 (0.05/5), 2-sided.

We assessed whether in lymphoma NHL and HL poor antibody response was a function of treatment related B cell depletion. Absolute lymphocyte count (ALC) was extracted from the EMR for all patients at the time-point immediately prior to vaccination. There was no correlation between ALC and response to vaccination across treatment groups, however mean ALC was significantly lower in antibody non-responsive NHL patients compared to antibody responsive NHL patients (p = 0.0246) (**Supplementary Figure 1**).

We further evaluated whether antibody titers increased between the first and second vaccination in the 18 patients who had time-points evaluable between the first and second vaccination. After the first vaccination only 3 of 18 (16%) of patients had a positive RBD. One patient (CLL on BTK inhibitor) increased their antibody titer after the second vaccination, however 17 of 18 patients did not have meaningful boost of their RBD immune titer after second vaccination (**Figure 2**). The absence of a substantial increase in antibody response after the second vaccination is in stark contrast to large increases seen in healthy volunteers in the clinical trial for the Pfizer vaccine (18).

**Figure 2.**
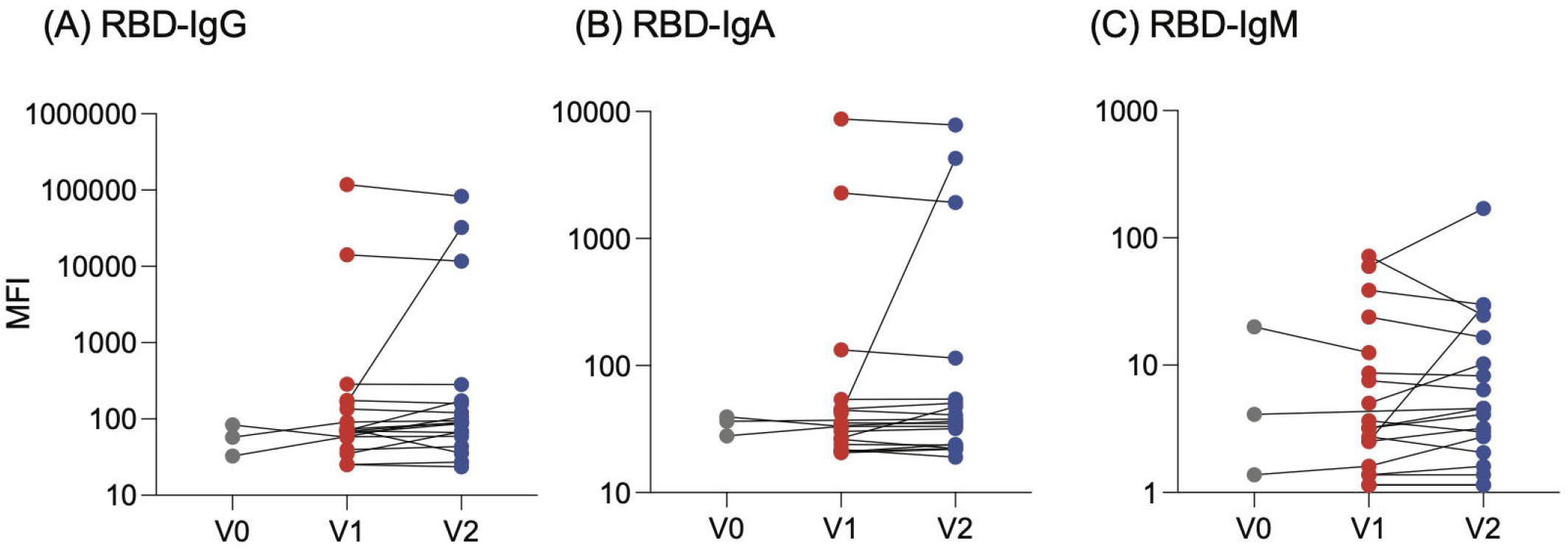
The levels of anti-Spike and anti-RBD IgG measured in MFI plotted versus the absolute lymphocyte count (ALC). ALC values are from the most recent blood work pre-vaccination.

### Impaired Functional Humoral Immunity in Antibody Positive Patients

To evaluate the functional humoral immune response, neutralizing antibodies were evaluated for antibody positive patients who demonstrated positive antibody titers. For patients receiving CD20 based therapy with or without chemotherapy, in 18 patients (75%), neutralizing antibodies were non-determined (ND) due to negative antibody response or insufficient antibody titers (**Figure 3**). For the 6 patients who were evaluable, 3 (13%) did not demonstrate any neutralization (NN) and only 3 (13%) demonstrated a neutralization IC50 ranging from 135 to 1767. In the NHL and CLL patients being monitored, 13 of 15 patients (87%) had neutralization ND; only 2 patients (13%) demonstrated neutralization antibody response with IC50s of 445 and > 10240. For 10 patients receiving BTK inhibitors, 4 (40%) were ND and an additional 2 patients (20%) were NN, leaving only 4 patients (40%) with active neutralization IC50 titers ranging from 33 to 1100. All HL patients were evaluable, and all patients had positive neutralization IC50s (43 to > 10240). When we correlated the neutralization titers with whole spike and RBD IgG we observed Regression R^2^= 0.42 and 0.61 to spike and RBD respectively, with a positive correlation of r = 0.65 and r = 0.78 respectively (**Figure 3**), similar to earlier reports on convalescent samples (14, 19). These data may suggest that patients who are able to mount a neutralizing antibody response could be protected from subsequent infection. However, a majority of patients who did mount a spike-specific antibody response had no neutralization or were non-determined, calling into question whether these patients are truly protected.

**Figure 3.**
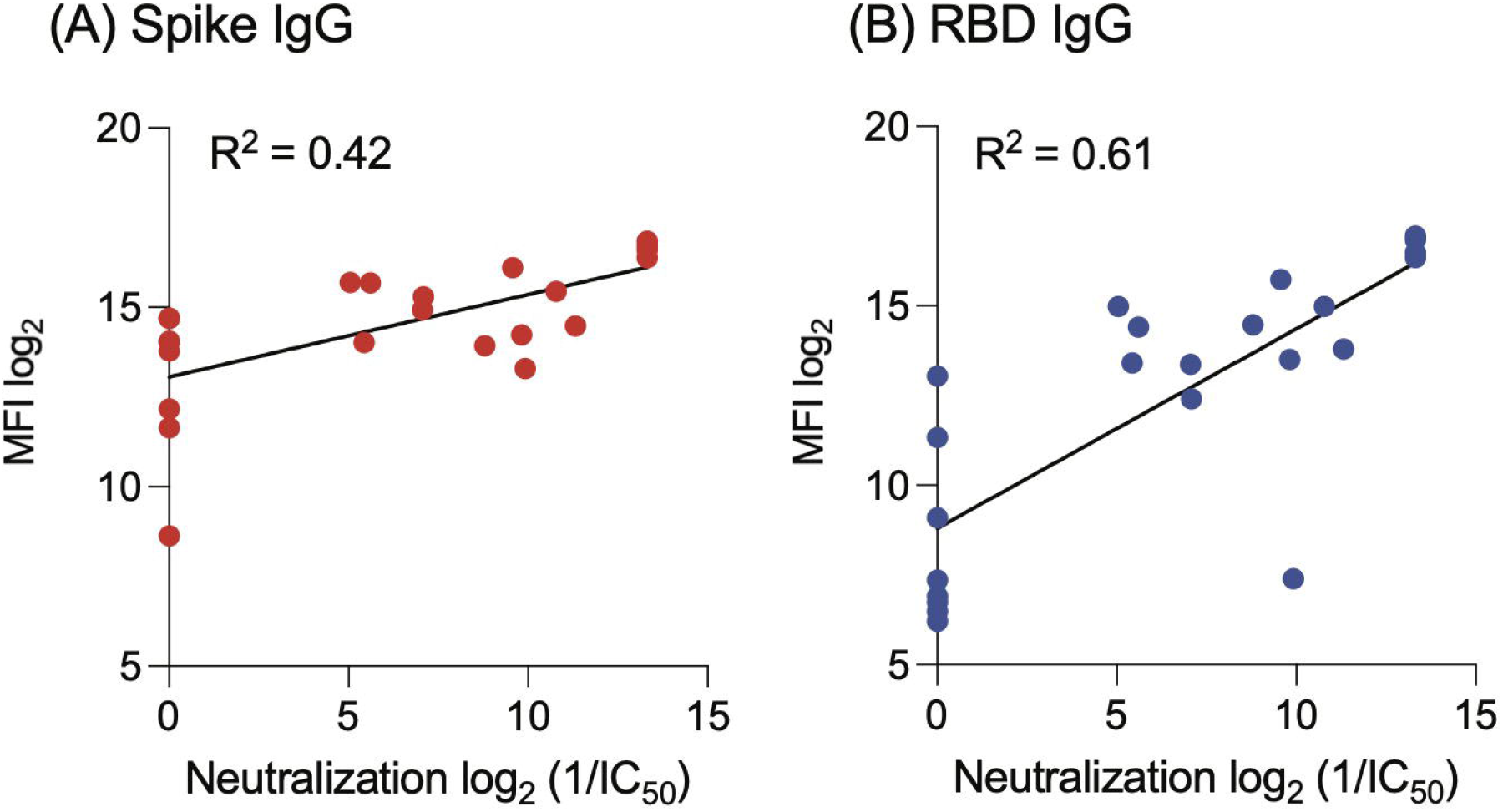
Correlation of the neutralization with the IgG levels. Anti-Spike (A) and anti-RBD (B) IgG levels measured in MFI are plotted against neutralization values. The lower limit of neutralization (13.32) were used for data points for which only the lower limit was determined. Regression R^2^= 0.61 and 0.42 for RBD and spike respectively was observed with corresponding correlation coefficients of 0.78 and 0.65 respectively.

## Discussion

Our study demonstrates that there is a decrease in the ability to mount an antibody response, as well as diminished antibody titers post SARS-CoV-2 vaccination in multiple NHL subtypes and CLL, especially in patients receiving anti-CD20 antibody therapy, and BTK targeted therapy. This decreased antibody response also occurred in patients monitored with an active tumor burden or more than 12 months from treatment (mean time from treatment 1671 days). Immune reactivity is lower for specific antigens for SARS-CoV-2 (the RBD) than for the whole spike protein, which may cross-react with antibodies from past exposure to other coronaviruses. This overall low response corresponds with previously reported data for NHL and CLL patients demonstrating a decreased immune response to influenza vaccines, and for CLL patients as has previously been reported to SARS-CoV-2 vaccination. However NHL and CLL patients demonstrated significantly lower antibody titers than normal controls, suggesting a qualitative as well as quantitative dysfunction in humoral immunity. More importantly, few antibody positive patients demonstrated function humoral immunity. Of the 24 patients receiving rituximab based therapy either alone or in combination with chemotherapy or other therapies, only 3 patients (13%) generated antibodies that neutralized authentic SARS-CoV-2 virus. The remaining patients either did not generate an antibody response, had antibody titers too low to assess for neutralizing antibodies, or did not produce neutralizing antibodies. These findings were irrespective of whether patients received one or both vaccinations, and which type of vaccine (Pfizer or Moderna) they received. Poor humoral immune function was not merely a function of treatment with rituximab or other B cell-targeting therapy; only 2 of 15 (13%) of patients (both NHL) who were being monitored generated neutralizing antibodies, and none of these patients had received therapy in over a year, most for many years. This suggests that the defects in humoral immunity observed in these patients are multifactorial, with significant disease-related factors in addition to the therapy effects mediating this immune deficiency.

Seventeen of eighteen patients in our study who had blood evaluable at 2 time-points failed to boost immunity after their second vaccination, which is in sharp contrast to the immunity described in normal volunteers vaccinated in the Pfizer and Moderna trials. This apparent inability to receive a boost from second vaccination speaks to profound dysfunction in humoral immunity and also raises the question of whether booster vaccinations provide any additional immune protection, or whether second vaccinations for these patients should be spaced further apart, as has been suggested with the Oxford AstraZeneca vaccine, where data supported a 12 week window between the first and second doses (20). This important question must be studied in larger patient populations to confirm this finding.

Responses to IgM and IgA were similarly lower to RBD IgM and IgA then to whole spike IgM and IgG for all patient groups. This suggests global humoral dysfunction across all arms of the humoral immune system, and may have implications for complement function, mucosal protection, and cellular immunity. Protective immunity to infection or vaccination is generated from combined activity of the humoral and cellular arms of the immune system. The primary defense against SARS-CoV-2 appears to be through the humoral arm of the immune system, supported by the clear benefit generated by treatment with monoclonal antibody therapy (21, 22), and the high antibody titers seen in convalescent patients (23). However, cooperation between humoral and cellular immunity has been suggested; CD4+ T cell response have been correlated with the degree of anti SARS-CoV-2 IgG and IgA titers (24), specificity of SARS-CoV-2 specific T cell response are significantly associated with a milder disease course (25, 26), and ineffective humoral immunity may underlie defects in neutralizing antibodies as these antibody responses are generally T cell dependent (27). The question of whether patients with inadequate humoral response to vaccination have similar dysfunction in cellular immunity is an important one and a high priority of future study.

Hodgkin lymphoma patients receiving chemotherapy did not demonstrate a significant impairment in the humoral response to vaccination. Data suggests that these patients have an impaired systemic immune response to recall antigens and to mitogens with their primary reported defect in cellular immunity, while their humoral immunity appears relatively unimpaired both from their disease and their cytotoxic chemotherapy (6, 28, 29). This suggests that B cell targeting therapies rather than conventional chemotherapy, may be the more important contributor to humoral immune dysfunction. Studies in solid tumor patients receiving chemotherapy may help to elucidate this question.

These data have important implications for timing of vaccination after B cell targeting therapy and public health recommendations, such as spacing of the two vaccine doses, and booster vaccinations for these patients. It is not clear from this data that postponing treatment for a few months to allow vaccination provides any protective benefit as all of our patients were within 6 months of treatment. It is an open question whether this defect persists for even longer, as impaired response to influenza vaccine and B cell depletion has been reported in patients receiving rituximab for up to one year (4). Whether these patients should be a priority for booster shots and whether further vaccination could augment humoral immunity are important questions that should be a high priority for investigation.

This data is of particular importance, given the recent guidance from the CDC that vaccinated patients no longer need to be masked indoors as well as outdoors. Patients with NHL or CLL who have active disease or who have been treated with either anti-CD20 antibody therapy or BTK inhibitors should not consider their immunity from vaccination to be assured. If infected with SARS-CoV-2, they should be a high priority for monoclonal antibody directed therapy. Unless immune response to vaccination is confirmed with laboratory testing, they should continue to mask, socially distance, and to avoid close contact with non-vaccinated individuals.

## Conclusion

Our data demonstrates that patients with NHL or CLL who have active disease or who are within 6 months of treatment with CD20 based therapy or BTK inhibitors demonstrate quantitative and functional defects in humoral immunity to SARS-CoV-2 vaccination, with less than 15% of patients producing neutralizing antibodies to the virus, and over 90% of patients failing to boost immunity after their second vaccination. Lymphoma and CLL patients receiving B cell targeting therapies should be counselled that they may not have sufficient immunity from the vaccine to consider themselves protected. Unless confirmed with laboratory testing, they should continue to mask, socially distance, and to avoid close contact with non-vaccinated individuals.

## Data Availability

Contact the corresponding author for original data

## Acknowledgements

This work was supported in part by the National Institutes of Health grants R21 AI158997, R01 CA194864 and R01 CA212608 (to SK) and P30CA016087 (to the Perlmutter Cancer Center), in addition to philanthropy funding to CD, KS and SK.

## Authorship Contribution

CD, MG, JG, and SK designed the study; CD and JC ran the study, CD, MG, BR, KH, TM, MK, DK, SF, and JuC contributed patients to the study, AK and SK performed the antibody testing, KS and ST performed the neutralization assays, CD, JC, AK, JG, KS, and SK analyzed the data and wrote the manuscript. All authors reviewed and approved the contents, and consented to publication of the current version of this manuscript.

## Conflicts of Interest

S.K. is a SAB member and holds equity in and receives consulting fees from Black Diamond Therapeutics and receives research funding from Puretech Health and Argenx BVBA. All other authors no relevant conflicts of interest.

## Notes

### Author Declarations

The Institutional Review Board of the Perlmutter Cancer Center at NYU Langone Health reviewed and approved the study, which was conducted according to the Declaration of Helsinki and International Harmonization Guidelines for Good Practice. All patients signed informed consent for the study.

