## Supplementary figures and images for "Impaired Humoral Immunity to SARS-CoV-2 Vaccination in Non-Hodgkin Lymphoma and CLL Patients"

### Supplemental Figure 1

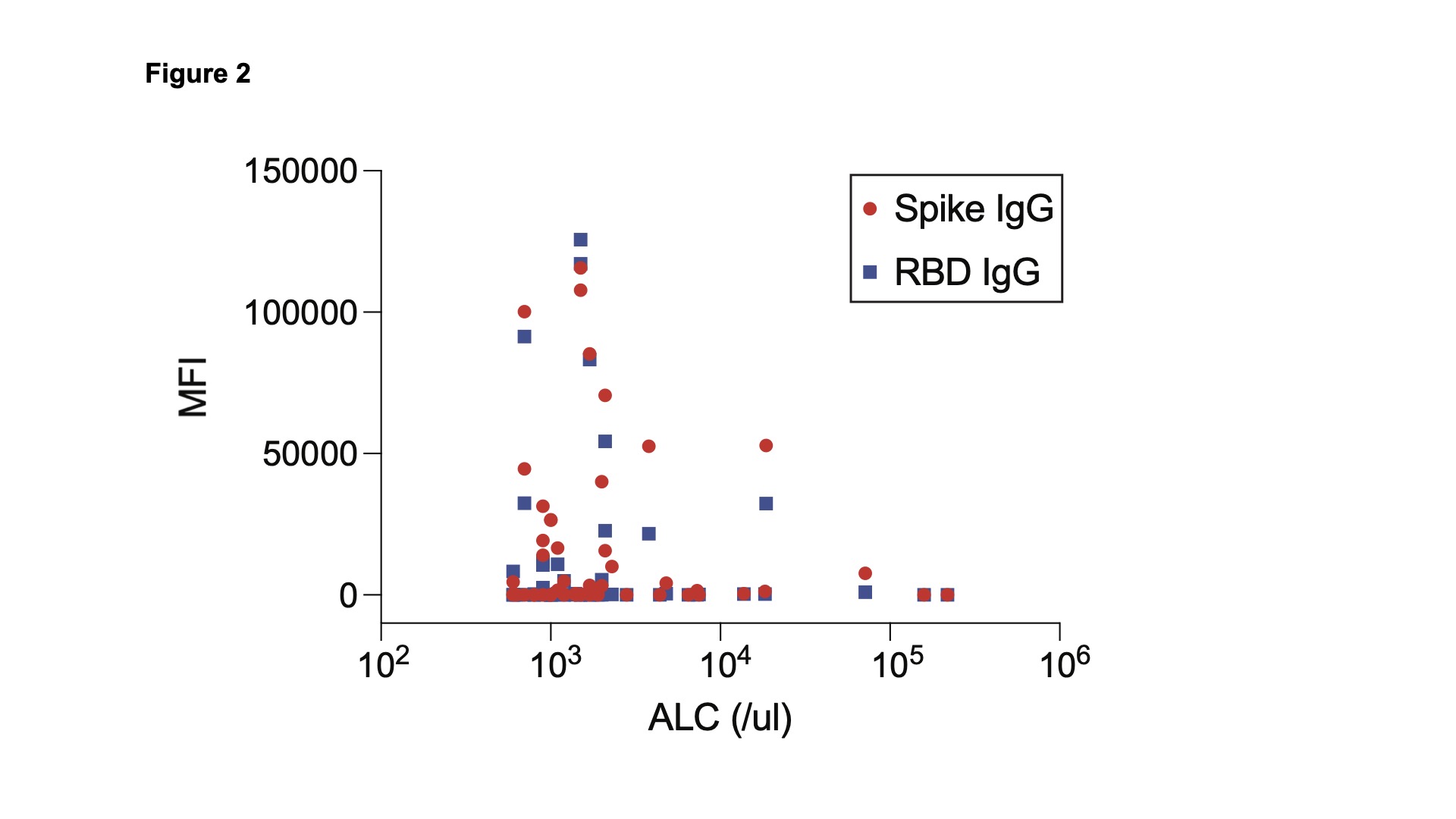
